# Intra-host evolution provides for continuous emergence of SARS-CoV-2 variants

**DOI:** 10.1101/2021.05.08.21256775

**Authors:** Justin T. Landis, Razia Moorad, Linda J. Pluta, Carolina Caro-Vegas, Ryan P McNamara, Anthony B. Eason, Aubrey Bailey, Femi Cleola S. Villamor, Angelica Juarez, Jason P. Wong, Brian Yang, Grant S. Broussard, Blossom Damania, Dirk P. Dittmer

**Affiliations:** Department of Microbiology and Immunology, The University of North Carolina at Chapel Hill School of Medicine, Chapel Hill, NC 27599, USA; Lineberger Comprehensive Cancer Center, Chapel Hill, NC 27599, USA; Kuopio Center for Gene and Cell Therapy, Kuopio, Finland

## Abstract

Variants of concern (VOC) in SARS-CoV-2 refer to viral genomes that differ significantly from the ancestor virus and that show the potential for higher transmissibility and/or worse clinical progression. VOC have the potential to disrupt ongoing public health measures and vaccine efforts. Yet, little is known regarding how frequently different viral variants emerge and under what circumstances. We report a longitudinal study to determine the degree of SARS-CoV-2 sequence evolution in 94 COVID-19 cases and to estimate the frequency at which highly diverse variants emerge. 2 cases accumulated ≥9 single-nucleotide variants (SNVs) over a two-week period and 1 case accumulated 23 SNVs over a three-week period, including three non-synonymous mutations in the Spike protein (D138H, E554D, D614G). We estimate that in 2% of COVID cases, viral variants with multiple mutations, including in the Spike glycoprotein, can become the dominant strains in as little as one month of persistent in patient virus replication. This suggests the continued local emergence of VOC independent of travel patterns. Surveillance by sequencing for (i) viremic COVID-19 patients, (ii) patients suspected of re-infection, and (iii) patients with diminished immune function may offer broad public health benefits.

## Introduction

The B.1.1.7 SARS-CoV-2 variant of concern (VOC) (20I/501Y.V1, S mutations N501Y, A570D, D614G, P681H, T716I, S982A, D1118H) arose because of long-term viral replication in an immunocompromised person in the UK ^1^. Several other case studies also attribute the emergenes of highly divergent variants to persistently infected patients ^2-7^ suggesting that intra-host evolution reflects a general mechanism for the continued emergence of highly divergent, potentially more transmissible SARS-CoV-2 variants. Each of these singular events has been linked to extraordinary clinical circumstances.

The rapid spread of the Spike protein D614G variant, which only had a single point mutation as compared to the earliest human isolate (SARS-CoV2/hu/CHN/Wuhan-Hu-1/2019), shows that novel SARS-CoV-2 variants can rapidly take over the population ^8-10^. Viral varients are more transmissible due to a variety of phenotypes, such as higher genome copy numbers in nasal secretions, increased environmental stability, or better / broader receptor utilization. Other mechanisms are possible as well, including the ability to maintain longer shedding periods, which we define as persistence, upon infection. Exactly, how frequent new VOC arise in persistently infected COVID-19 patients is unknown.

VOC refer to viral genome sequences that differ dramatically from their most common recorded ancestor, typically by ≥3 single nucleotide variations (SNVs), typically with non-synonymous mutations in the Spike (S) glycoprotein. In the literature, VOC display increased infectivity in tissue culture, increased human-to-human transmission patterns, and worse clinical outcome. VOC are predicted to have decreased vaccine-induced neutralization and to be resistant to therapy using monoclonal antibodies. As, VOC have the potential to render ongoing public health measures and vaccine efforts ineffective it is important to identify situations that foster the emergence of VOC.

In addition to B.1.1.7 (20I/501Y.V1) first recognized in the UK, the US CDC lists B.1.351 (20H/501.V2, S mutations: K417N, E484K, N501Y, D614G, A701V), first recognized in South Africa, and B.1.1.28.1 (P1/20J/501Y.V3, S mutations: L18F, T20N, P26S, D128Y, R190S, K417T, E484K, N501Y, D614G, H655Y, T1027I) first reported in Brazil as VOC. Sequencing surveys identify new VOCs almost daily such as B.1.429 and B.1.427 (CAL.20C, S mutations: S13I, W152C, L452R, D614G or S mutation: L452R), which are rapidly expanding and are associated with worse clinical outcome ^11^.

An open question in the field is whether VOC emergence represents a singular, low frequency event in time and space ^12^, in which case travel restrictions would be an appropriate measure of containment, or if VOC continuously and repeatedly emerge in local communities. In the latter case, surveillance by whole genome sequencing would be an urgent and prudent course of action. This study estimates that approximately 2% of persistently infected COVID patients develop highly divergent variants in as little as three weeks.

## Results

We identified n=94 cases of COVID-19 with 2 or more positive SARS-CoV-2 tests (**Figure 1A**). The case definition included cases where intermittent viral load assays were negative, as it is not possible, a priori, to distinguish between persistent replication below the level of detection and independent reinfection. The median age was 52.6±17.4 years (mean ± sd); 3 participants were under the age of 18. 40/94 (43%) of participants were female. The cases covered the period from April 1, 2020, to October 17, 2020. During that time the COVID-19 epidemic was accelerating in the local community and was primarily driven by symptomatic transmission events ^13^. As most of the data were from de-identified patient records, the clinical presentations of each case could not be conclusively established. The inclusion criterion was solely based on viral detection (positive/negative) by CLIA assay in two consecutive NP swabs.

**Figure 1:**
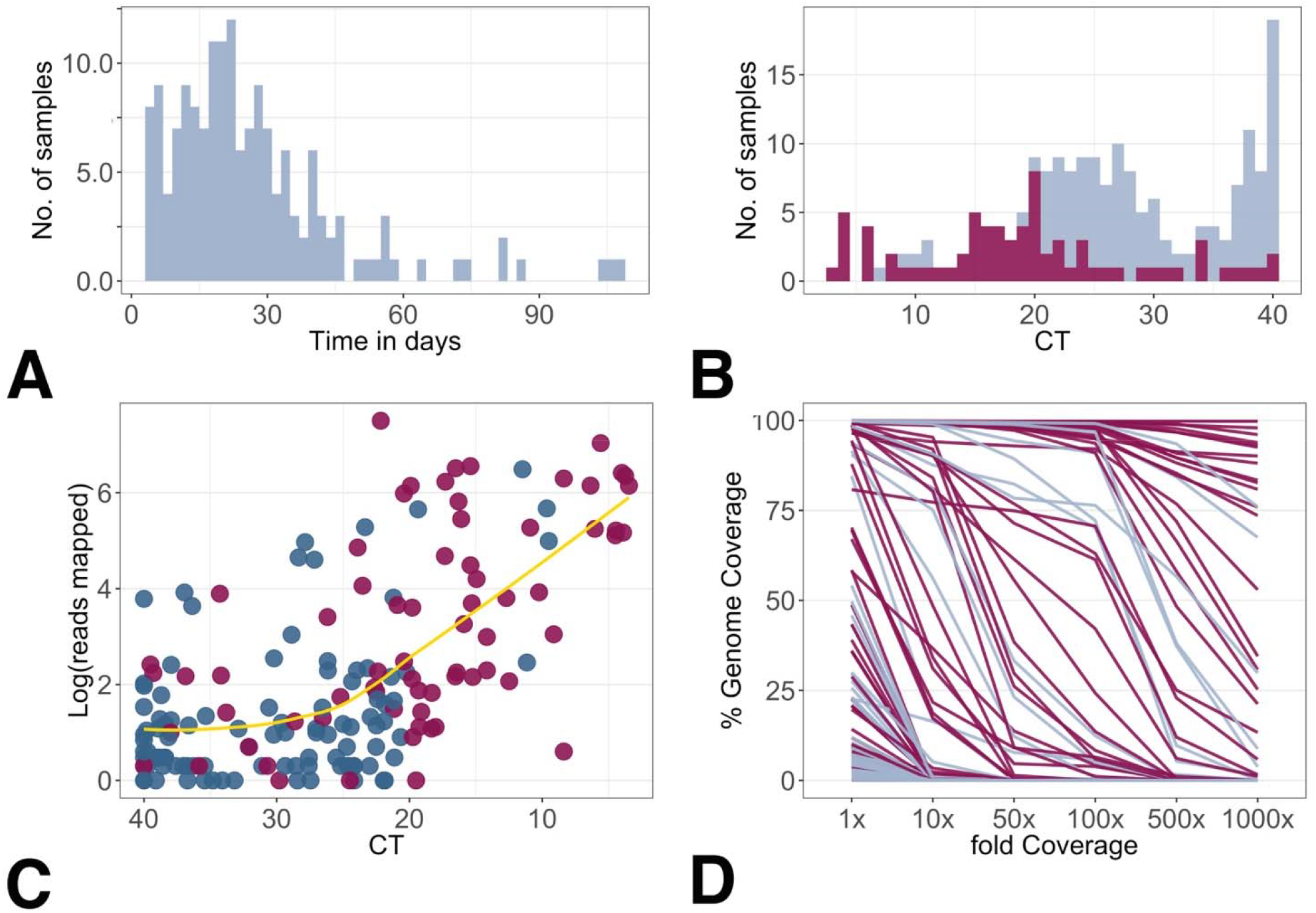
Summary characteristics of the cohort and sequencing performance. Red indicates values at baseline (T0), blue indicates subsequent sampling points. (A) Distribution of observation time for the cohort. (B) Distribution of relative viral genome copy number as determined by real-time RT-QPCR. This excludes n=60 samples for which no CT values were available, only a negative/ positive determination. (C) Relation between genome copy number as determined by real-time RT-QPCR and fraction of reads mapped/total reads. (D) Relation between the fraction of reads mapped/total reads and coverage at 1x, 10x, 100x.

All participants had detectable viral load at baseline. At subsequent time points, relative log_2_ viral genome copy number, as measured in RT-QPCR cycle number threshold (CT), declined in most cases as indicated by higher CT values (**Figure 1B**). In many samples, genome copy numbers at late time points were near the limit of detection (CT>30). This was expected since even during acute symptomatic infection with SARS-CoV-2, genome copy numbers peak during a few days over the course of infection. All samples were subjected to targeted amplification of SARS-CoV-2 and next-generation sequencing (NGS) as described previously ^8^.

Most participants did not have high enough viral RNA levels at time points subsequent to baseline to generate sufficient reads to yield complete genome-wide coverage at the required sequencing depth. Nevertheless, NGS conclusively confirmed the presence of SARS-CoV-2 RNA in the real-time RT-QPCR positive samples and in none of the RT-QPCR negative samples. The number of mapped reads showed a log-linear relationship to CT (**Figure 1C**) for CT values ≤ 24.71 (^95^CI: 19.11-33.60) and was uncorrelated for samples with CT > 24.71. As expected. more mapped reads correlated with higher overall genome coverage (**Figure 1D**). These performance characteristics are in line with other studies.

To ascertain the linear range for SNV calling a dilution series of SARS-CoV-2/hu/USA/WA1/2020 was generated (**Table 1**). Only single nucleotide variants (SNV) with QUAL scores > 200, as called by samtools ^14^, were included in subsequent analyses. The SARS-CoV-2/hu/USA/WA1/2020 stock has 8 SNVs as compared to SARS-CoV2/hu/CHN/Wuhan-Hu-1/2019 (NC_045512). These 8 SNVs were recovered over a dilution range of 4 log_10_ orders of magnitude down to a limit of detection of 7 pfu/ml. The experiment was repeated with an artificial RNA substrate carrying 428 single point mutations and the same dilution range. From this, 423 single-point mutations were consistently sequenced over the entire dilution range. This experiment demonstrates that the limit of detection for NGS-based SNV typing is at or below the detection limit of viral culture. By implication every variant sequence reported here represents infectious virus, rather than residual fragments of viral RNA.

**Table 1:** SNV calling sensitivity. SNV refers to the number of correctly called SNVs relative to SARS-CoV2/hu/CHN/Wuhan-Hu-1/2019 (NC_045512) for either an artificial RNA with 428 point mutations or WT virus strain 2019-nCoV/USA-WA1/2020. Dilution factor shows the dilution. IT refers to infectious titer in plaque forming units/ ml. Coverage revers to coverage at 1x. The raw reads are available as bioproject PRJNA719089.

| Sample | Dilution Factor | IT in pfu/ml | Coverage | SNV |
| --- | --- | --- | --- | --- |
| MT RNA (n:428) | 1 | NA | 100.00% | 426 |
| . | 4 | NA | 100.00% | 427 |
| . | 16 | NA | 100.00% | 427 |
| . | 64 | NA | 100.00% | 426 |
| . | 256 | NA | 100.00% | 427 |
| . | 1,024 | NA | 100.00% | 428 |
| . | 4,096 | NA | 99.84% | 423 |
| . | 16,384 | NA | 98.27% | 423 |
| WT virus (n: 8) | 1 | 115,000 | 100.00% | 8 |
| . | 4 | 28,750 | 100.00% | 8 |
| . | 16 | 7,188 | 100.00% | 8 |
| . | 64 | 1,797 | 100.00% | 8 |
| . | 256 | 449 | 100.00% | 8 |
| . | 1,024 | 112 | 100.00% | 8 |
| . | 4,096 | 28 | 100.00% | 8 |
| . | 16,384 | 7 | 100.00% | 8 |

Sequence diversity in the data set is based on individual SNV that could be ascertained with high confidence. A total of n = 882 SNVs passed the QC filters. **Figure 2A** shows the average number of SNVs for each sample (n=86). The mode was 11, two samples had n≥20 and n≥40 SNVs, respectively, which indicate a highly divergent viral variant. The majority of SNVs detected had >90% frequency in the sample (**Figure 2B**), although a subset of samples also had SNVs with lower frequency. SNVs with <50% frequency were excluded from the analyses presented here, although minority variants down to 10% frequency were reliably identified. Hence, pair-wise comparisons between samples represent a lower limit of sequence divergence over time. **Figure 2C** depicts the counts of non-synonymous SNVs for each SARS-CoV-2 gene across all samples. As expected total SNV counts correlate with the gene size. Also indicated are the SNV frequencies for each gene.

**Figure 2:**
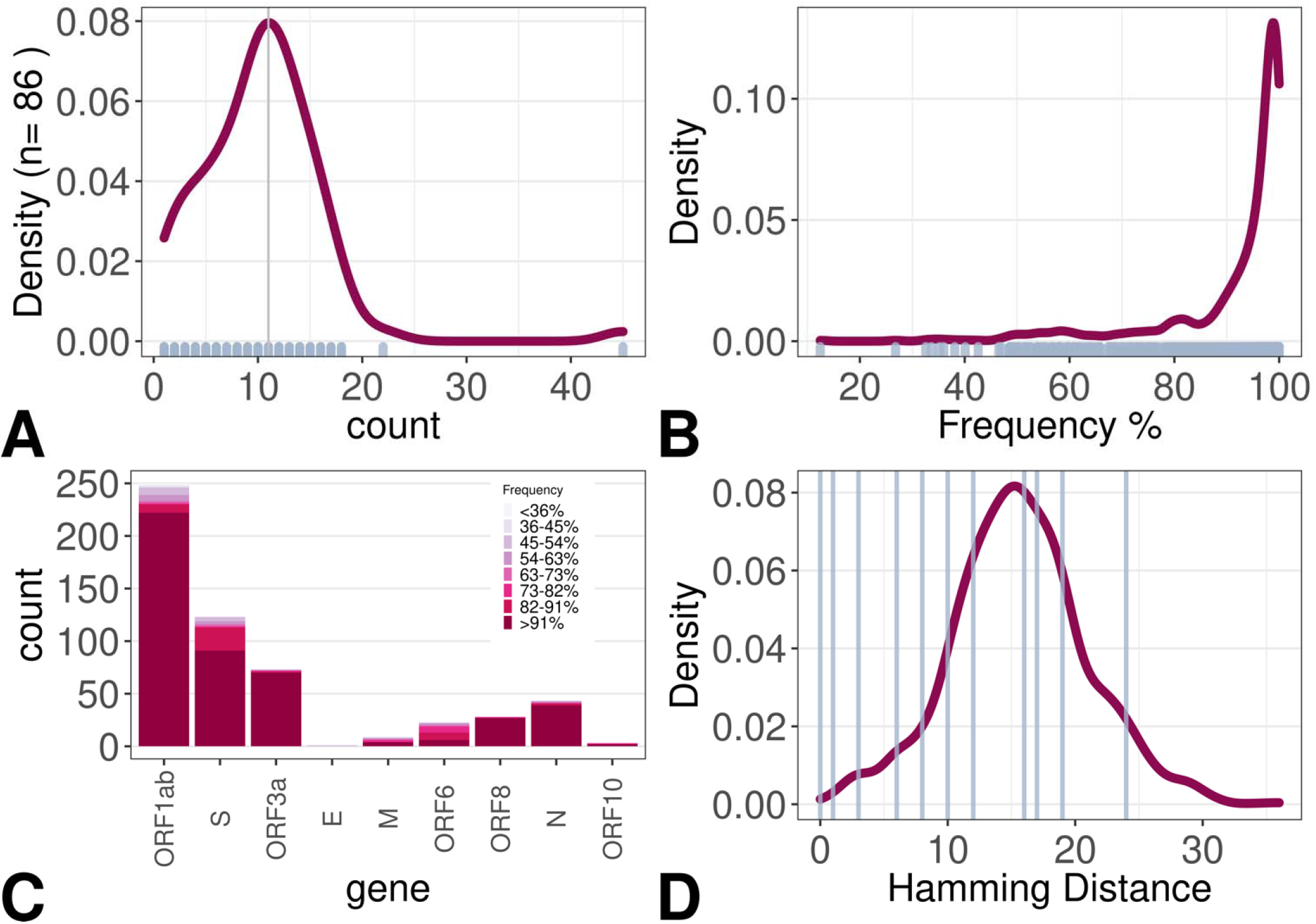
Summary of genome diversity. (A) Distribution of SNVs per sample. (B) Distribution of SNV frequency in the data set. (C) Count of non-synonymous SNVs per gene, color-coded by frequency of the SNV (D) Distribution of Hamming distances in the data set. Blue lines indicated paired samples.

It is not possible, a priori, to decide whether a new SNV is the result of de novo mutation during persistence in the host or selection of a strain that was present at with a frequency below the limit of detection at the initial time point of infection. **Figure 2D** shows the empirical density distribution of Hamming distances among all samples. The Hamming distance is defined as the minimum number of substitutions between any two sequences. It is less or equal to the genetic distance and not dependent on the time or the rate of evolution. Hence, it represents a lower limit on genetic diversity. Paired samples, i.e., samples from participants with two consecutive timepoints that each yielded enough material for sequencing are indicated by blue lines. The majority of pairs had smaller than mean Hamming distances, consistent with the hypothesis that intra-host replication accumulates fewer mutations than inter-host transmission. Two paired samples had Hamming distances that were larger than the mode, indicative of accelerated intra-host evolution during persistence or a second, independent re-infection event.

The null hypothesis stipulates that upon infection SARS-CoV-2 replicates rapidly, synchronously, and without accumulating mutations due to the intrinsic low error rate of RNA-dependent RNA polymerases of the Coronaviridae ^15^ as well as limited host selection pressure prior to the onset of adaptive immunity. Most symptomatic COVID-19 cases are viremic for two weeks or less ^16,17^. Consistent with this hypothesis, 4 pairs of participant sequences were identified with ≤ 1 SNV difference across the entire genome. These represent the prototypical infection scenario for coronaviruses.

Other cases accumulated more SNVs. **Figure 3A** depicts a phylogenetic tree obtained from the multiple alignments of n=48 completely-sequenced SARS-CoV-2 genomes (data available at GISAID). In addition to samples from this study the first two SARS-CoV-2 confirmed cases in the catchment area were included: hCoV-19/USA/NC-CDC-6999/2020 from March 3, 2020 (2020-03-03), which is a Spike (S) 614D variant and hCoV-19/USA/NC-CDC-0034/2020 from March 8, 2020 (2020-03-08), which is an S 614G variant. The reference genome SARS-CoV-2 SARS-CoV2/hu/CHN/Wuhan-Hu-1/2019 was set as root. The tree was generated from a MAFFT alignment further processed by MrBayes (HKY85 substitution model with unconstrained branch length using SARS-CoV-2 SARS-CoV2/hu/CHN/Wuhan-Hu-1/2019 as outgroup). The monophyletic sequence pairs in blue (LCCC0230/LCCC0235, LCCC0246/LCCC0247, LCCC0220/LCCC0224, LCCC0192/ LCCC0228) conform to the null hypothesis of limited intra-host evolution. These pairs accumulated ≤1 SNV over a two-week period (range: 7-19 days) and represent stable, canonical infection events.

**Figure 3:**
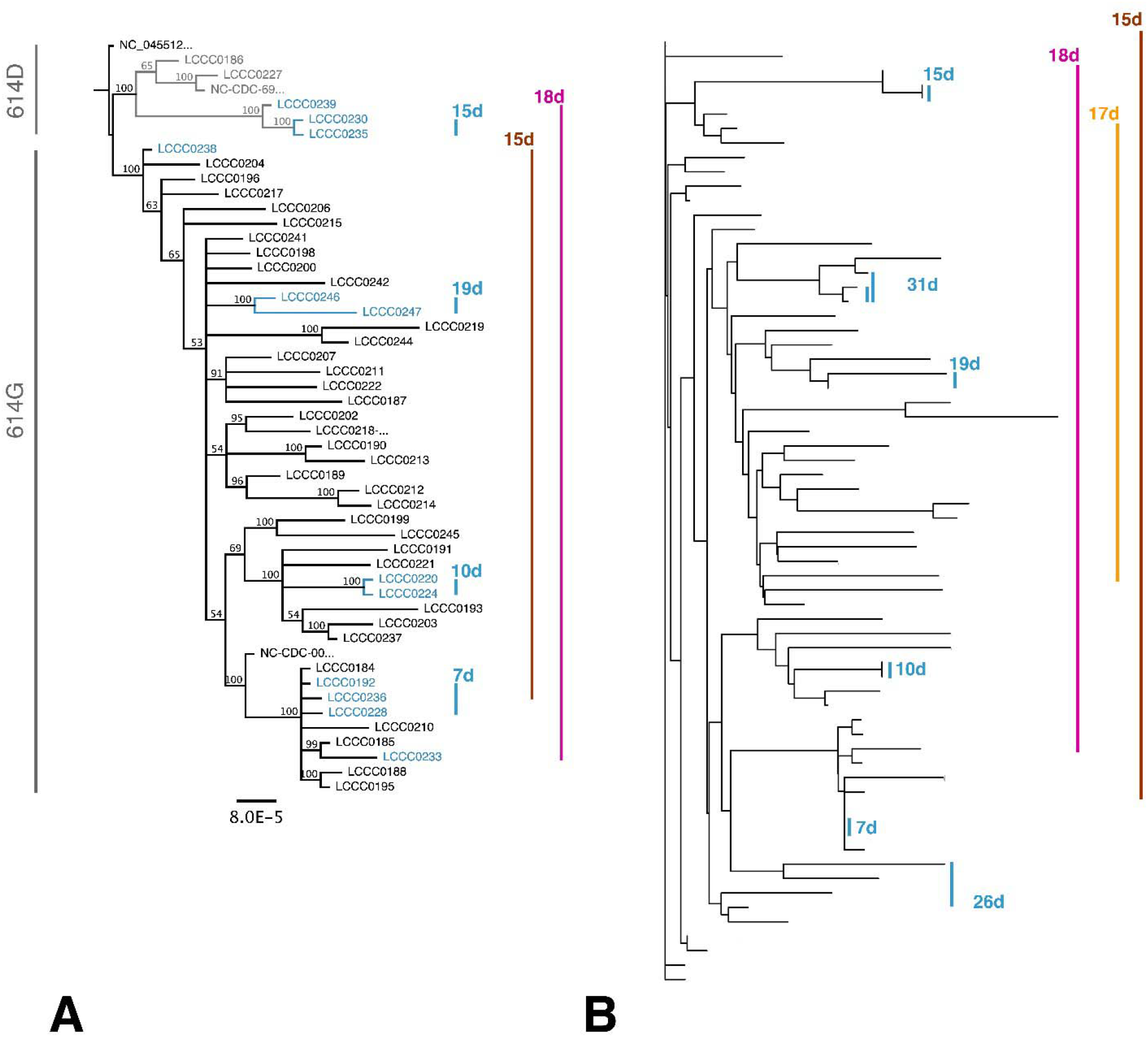
(A) Phylogenetic tree obtained from a multiple alignment of n=48 whole SARS-CoV-2 genomes as submitted to GISAID. Tree under the assumption of unequal evolution rates. Paired samples are indicated by colored lines and labeled by distances in days. (B) Phylogenetic tree based on n = 231 high quality SNV position for n= 67 complete and partial SARS-CoV-2 genomes. Paired samples are indicated by colored lines and labeled by distances in days.

The two sequence pairs LCCC0233/ LCCC0239 and LCCC0238/ LCCC236 represent cases where 9 SNVs accumulated over 15 days and 23 SNVs over 18 days, respectively. This resulted in the sequences being distant on the phylogenetic tree. LCCC0233/ LCCC0239 differed by the presence of the S D614G mutation that is associated with high transmissibility and biological fitness in culture ^8-10,18,19^. Their pattern was inconsistent with the null hypothesis and supports scenarios of either accelerated intra-host evolution or independent superinfection.

It was difficult to obtain high-quality, whole-genome sequences from samples with low genome copy numbers. To expand the set of paired samples available for analysis, additional genomes were analyzed even if they were incomplete (> 1000 N at 1x coverage). This was possible because even for incompletely covered genomes, individual SNVs, relative to the reference, could be ascertained with high confidence (QUAL scores > 200). This yielded an extended pool of n=67 samples, including the prior samples with complete genomes, which were used to construct a Hamming-distance matrix. **Figure 3B** shows a neighbor-joining tree based on this matrix and rooted at SARS-CoV2/hu/CHN/Wuhan-Hu-1/2019. The clustering by informative SNVs alone was consistent with the clustering based on analysis of the entire viral genome sequences as expected. The expanded data identified 6 paired samples, which diverged only minimally from each other during the observation period of 7, 10, 15, 19, 26, 31 days, respectively. By contrast, 3 sample pairs that were not consistent with the null hypothesis of limited intra-host evolution. These three pairs represent SARS-CoV-2 strains that have diverged significantly from the initial isolate over a period of 15, 17, and 18 days, respectively. Note how the observation periods for the two groups overlap: over a time range of 7-31 days, the majority of patients accumulated ≤1 SNV, a minority accumulated ≥ 9 SNV in the SARS-CoV-2 genome. This demonstrates that the conditions under which VOC could evolve are present in a significant fraction of COVID-19 patients with detectable viral loads at greater two weeks past infection.

To put these samples in the context of the local sequence diversity, an additional 226 whole-genome sequences that were generated from the same participant pool by the same technology and bioinformatic pipeline were assembled and a timed consensus tree generated using BEAST (**Figure 4**). The result was consistent with exponential growth during the observation period. The SARS-CoV2/hu/CHN/Wuhan-Hu-1/2019 genome was set as root. The two initial isolates in the state of North Carolina, one representing the D614 clade (SARS-CoV-2/human/USA/NC-CDC-6999/2020) and the other the G614 clade (SARS-CoV-2/human/USA/NC-CDC-0034/2020) were added for orientation. The resultant phylogram again identified the same four pairs as before, representing minimal intra-host evolution as well as the two pairs that did not cluster together and represent accelerated intra-host evolution. The paired samples belonged to different lineages (PANGO classification), as evidenced by their distribution among the other samples. This suggests that different evolutionary speeds are not the property of a rare mutator varient of SARS-CoV-2 but represent independent events that took place repeatedly.

**Figure 4:**
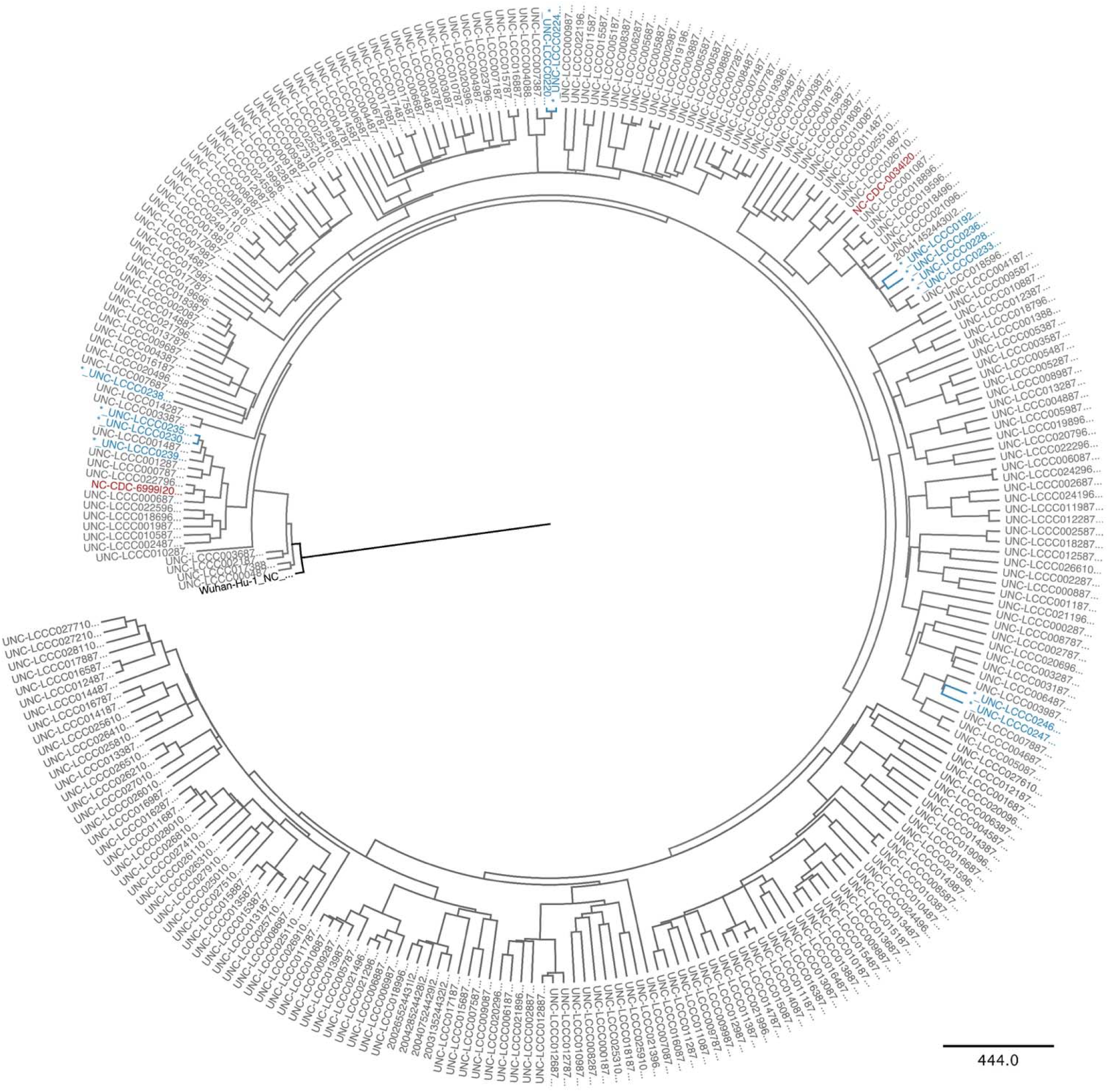
Phylogenetic tree obtained using BEAST after a multiple alignment of n=273 whole SARS-CoV-2 genomes from GISAID that were collected at the same time in the same population and determined by the same technology and bioinformatics pipeline. The initial introductory events are indicated in red, the root Hu-Wu-1 in black, and paired specimen in blue.

Two scenarios could account for these observations. First, in some patients, SARS-CoV-2 evolution and mutation rates were accelerated as compared to rates determined by molecular clock phylogenies of inter-host events. This scenario also includes the sequential emergence and/or disappearance of dominant subpopulations. Second, an independent infection event, i.e., superinfection, took place prior to the last sampled time point. Sequence analysis alone cannot distinguish between these two scenarios. Assuming that the patients were in isolation during hospitalization and followed CDC guidelines regarding quarantine and masking for patients who have recovered from COVID-19, a superinfection event seemed less likely than persistent infection, but it remains a formal possibility. In sum, this study supports the idea of intra-host evolution during the persistence of SARS-CoV-2 in a significant number of infections that can server as a reservoir for the continued emergence of highly divergent variants, including VOC.

## Discussion

The SARS-CoV-2 epidemic in NC was seeded by two singular events, one introducing the D614 variant strain on March 03, 2020, and another event introducing the G614 variant strain on March 13, 2020 ^8^. Since then, we surveyed SARS-CoV-2 infected persons by NGS. This cohort identified 94 patients who were SARS-CoV-2 positive on at least two occasions as determined by real-time RT-QPCR under CLIA-compliant diagnostic testing. 6 patients had ≤1 SNV difference for samples drawn approximately 14 days apart. 3 patients had accumulated ≥1 SNV during the same time period. One participant had accumulated 23 SNV over a 19-day period. The three participants with extensive viral evolution had potentially comorbid conditions: one participant had a history of infections, another had diabetes mellitus.

First, these data demonstrate that the majority of SARS-CoV-2 infections follows a paradigm of limited intra-host viral evolution consistent with the absence of selection in a naïve host and are consistent with the reported mutation rate of ∼ 1 SNV per genome per 14 days ^2,20-22^.

Second, these data demonstrate that reinfection is a possibility in persons previously exposed to SARS-CoV-2, which is consistent with an ever-increasing number of case studies ^23-28^. It is curious that the one potential reinfection case with the largest numbers of SNV displayed the earliest isolate carrying the S D614 SNV and the second infection was with the more transmissible and infectious S G614 variant, akin to the report by To et al. ^26^. We speculate that natural infection with one receptor-binding domain (RBD) variant may not protect extremely well against infection with an RBD second variant, even though vaccination does ^29,30^. This finding affirms the recommendation for universal vaccination even for persons previously exposed to SARS-CoV-2.

Third, these data demonstrate that variants with a large number of SNVs regularly emerge in persons that carry SARS-CoV-2 as early as two weeks after primary exposure. Accelerated evolution has been described for coronaviruses, e.g., those carrying a mutator polymerase or exposed to a mutating drug ^15^. Accelerated evolution can also be achieved by sequential bottlenecks, such as generated during persistent low-level infection where the host immune responses dramatically decimate, but never fully eradicate the virus ^25,31,32^. Even under the conservative assumptions of (a) the one most divergent sample in our study being due to reinfection with an unrelated strain and (b) the other persistently positive patients in the cohort, for which a second genome sample was incomplete, having no SNVs, we estimate that highly divergent variants of SARS-CoV-2 emerge at a frequency of 2/94 (2%) among hospitalized COVID-19 patients.

There are limitations to this study. First, it is a single center study biased towards patients with severe clinical disease rather than a population-based sample. This contrasts with the majority of SARS-CoV-2 infected persons, who do not require hospitalization. Second, many participants did not have enough viral RNA at time points subsequent to baseline for NGS to yield complete genome coverage. This was expected as in most cases, viral RNA is detectable by swab of the nasopharyngeal cavity (NP-swab) 2-3 days before onset of symptoms (pre-symptomatic), but rapidly disappears within 5 days after clinical symptom onset^17,33-36^. The genome copy numbers reported here are consistent with a report of 35 RT-QPCR positive specimens that were collected at >10 days after symptom onset, and that failed to yield infectious virus upon culture ^25,37,38^. Control experiments established that complete genome coverage and confident SNV detection was possible down to a limit of 7 pfu/ml; however, it is possible that persistent intra-host replication predominanty leads to the accumulation of fragmented viral genomes or debilitated viral particles with low transmission potential as the intra-host selection pressures differ from the selection pressures that lead to increased transmissibility between hosts.

In sum, this study suggests that widely divergent SARS-CoV-2 variants, including VOC, will continually emerge spontaneously as long as there is significant community infection (often stipulated as above 100/100.000 cases over a 1 week period). Intensified, repeat monitoring by sequencing of hospitalized COVID-19 patients and infected at-risk persons, such as persons on immunosuppression or cancer chemotherapy ^32^, is useful in identifying new VOC. It may have direct clinical as well as public health benefits. This study also supports the notion that wide-scale vaccination efforts are needed at a global level to lower the spread of SARS-CoV-2 in the human population and thus prevent the emergence of new VOC.

## Data Availability

Full-length SARS-CoV-2 genomes that met the confidence and quality criteria detailed below were uploaded to GISAID. Other sequences, including index cases, were obtained through GISAID.

## Acknowledgments

We thank the McLendon Clinical Laboratories for providing de-identified remnant samples and helpful discussions, specifically Melissa Miller and Shawn Hawken, as well as the UNC department of medicine, Division of Pulmonary Diseases, and Critical Care Medicine for providing de-identified clinical data, specifically William Fischer, and Subhashini Sellers. We thank AccuGenomics Inc., for providing material for beta testing. This work was supported by funding from the Medical Foundation of North Carolina, the NC Collaboratory, and public health service grants 5UM1CA121947-10 to R.P.M, 2P01CA01901438 to J.D.G, and B.D., and 2-R01-DE018304-10, 1-R01-CA239583-01 to D.P.D.

## Author Contributions

Conceptualization, B.D., D.P.D; Methodology R.M., L.J.P., C.C-V. R.P.M, Software, J.T.L., A.B., B.Y.; Investigation, J.T.L, R.M, L.J.P., C. C-V, R.P.M, A.B.E, FC. S. V., A.J., J.P.W., G.S.B.; Writing – Original Draft, J.T. L, R.M., D.P.D; Writing – Review and Editing, B.D., D.P.D., A.B.E.; Visualization, J.T.L., R.M., D.P.D., Supervision, D.P.D., Project Administration, D.P.D., B.D.; Funding Acquisition, D.P., B.D.

## Competing interests

Authors declare no competing interests.

## Methods

### Resource Availability

#### Lead contact

Further information and requests for resources and reagents should be directed to and will be fulfilled by Dirk Dittmer.

#### Materials Availability

Full-length SARS-CoV-2 genomes that met the confidence and quality criteria detailed below were uploaded to GISAID. Other sequences, including index cases were obtained through GISAID.

#### Data and Code Availability

All sequence mapping algorithms and codes are publicly accessible, are elaborated in detail below, or available using the CLC Genomics Workbench V 2.0 (QIAGEN). R code that was used for data analysis is located on an accessible bit bucket folder https://ddittmer@bitbucket.org/dittmerlab/intermittent_covid_unc.git. Alignments, analyses, and statistical groups were made as previously described ^8^.

### Experimental Model and Subject Details

#### Sample Collection and Deidentification

This study used remnant samples of universal transport media (UTM) from provider-collected deep nasopharyngeal (NP) swabs after their clinical purpose had been completed. The SARS-CoV-2 status of each sample was determined at The University of North Carolina at Chapel Hill Medical Center (UNCMC) McLendon Clinical laboratories. None of the samples carried any identifiers other than the date of testing, age, and sex. Sample use was approved under human subjects approvals # 20-2448 and # 13-2140 by the Institutional Review Board (IRB) at the University of North Carolina; CB 7097, 720 Martin Luther King Jr. Blvd. Bldg # 385, Second Floor, Chapel Hill, NC 27599-7097.

### Method Detail

#### RNA isolation

RNA was isolated using a Roche Magnapure 24 instrument and kits according to the manufacturer’s protocol. In brief, 200 µL of UTM were neutralized with the addition of 0.1% Triton X-100 (proteomics grade, VWR: 97063-864) and 1X phosphate-buffered saline (PBS, Life Technologies, Catalog # 14190-144) to a final volume of 1.0 mL. Samples were incubated at room temperature for 30 minutes in a barcoded 2.0 mL screw cap tube (Roche, 07857551001) vortexing every 5 minutes for 15-second pulses. The solution was then processed through the Magnapure 24 instrument using the total nucleic acid extraction protocol. The MagNA Pure 24 Total NA Isolation kit was used for RNA extraction (Roche, 07658036001). Carrier RNA (Macherey-Nagel, 740514) was added to a final concentration of 9 ng/µL. For each processing batch, a negative reagent control and a negative cell pellet control were used. The reagent control consisted of 250 µL of 1X PBS instead of 250 µL of the sample in UTM. The 100 µL of purified RNA was processed for sequencing and viral load as described below.

#### Real-time qPCR

Relative viral genome copy number and cycle thresholds (Ct) was ascertained by real-time qPCR using primers and procedures previously published ^39^ and a protocol previously described ^8^. In brief, 30 µL input RNA was subjected to random hexamer-primed reverse transcription using the High-Capacity cDNA Reverse Transcription Kit (Applied Biosystems, 4368814). 9 µl cDNA was used for qPCR containing 125 nM for each primer and SYBR green as the method of detection on a Roche LC480II Lightcycler and Ct values determined by an automated threshold method.

#### Next-Generation Sequencing

Amplicon-based next-generation sequencing was performed using the Thermo SARS-CoV-2 Ampliseq kit. We used Genomic RNA from SARS-CoV-2, Isolate USA-WA1/2020, as a positive control (BEI Resources, NIAID, NIH: NR-52285). All samples were sequenced using random hexamer/oligo dT priming according to the manufacturer’s protocol on an Ion Torrent Chef (ThermoFisher 4484177) using Ion S5 Chef Solutions (ThermoFisher A27754). Samples were then loaded onto the IonTorrent S5 sequencer (ThermoFisher A27212) equipped with 530 Chip (ThermoFisher A27763). The amplicons are tightly tiled and overlapping. Amplicon sizes ranged between 68 and 232 nucleotides after trimming of low-quality sequences (Q20) and primer sequences (125-275 before trimming).

#### Bioinformatic analysis

Following primer trimming according to the manufacturer’s recommendations, additional, custom steps were added. Specifically, all sequences were quality trimmed using the bbduk script (arguments: qtrim=rl trimq=20 maq=20 minlen=40 tpe tbo) from bbmap version 37.36. The trimmed reads were mapped to the SARS-CoV-2 reference sequence (Accession: NC_045512) using bbmap. From each mapping, the following was collected: sequence variants, mapping coverage, and a consensus sequence. Sequence variants were called from the mapping file using samtools and bcftools ^14^. Mapping coverage was generated using ‘Deeptools’ bamCoverage ^40^. Sequence variants and mapping coverage were used to build the consensus sequences using bcftools. Only variants with a reported QUAL greater than 200 were included in the consensus and any region with 0x coverage were masked with Ns inserted for ambiguity. All consensus sequences derived from this study were curated to revert poly-nucleotide-tract mutations to the reference sequence. Lineages were assigned using Pangolin v.2.0 ^41^. Complete genomes have been submitted to GISAID and raw reads to SRA archives. Nomenclature as per International Committee on Taxonomy of Viruses (ICTV) ^42^.

#### Phylogenetic reconstruction

The alignments of complete genomes for this study were performed using MAFFT ^43^ and the initial phylogenetic tree based on whole viral genomes were generated using MrBayes ^44^ or RaXMAL ^45^ as implemented in Geneious Prime® 2021.0.3. using the HKY85 substitution model and gamma-distribution-based nucleotide rate variation with unequal branch lengths. NC_045512 was used as an outgroup.

For alignment of all NC sequences as obtained from GISAID, MAFFT, and RAxML or FastTree was used for initial alignment. The alignment was exported and used as input for a time-scaled Bayesian Tree was generated using BEAST v1.10.4, with the BEAGLE v3.1.0 library program ^46^. Estimated base frequencies using the Gamma distribution site model was selected. For the tree prior, the coalescent exponential growth rate was selected, using a random starting tree and a strict molecular clock. The Markov Chain Monte Carlo (MCMC) chain length was set to 10,000,000 steps sampling after every 1000 steps (5-7). Trees were annotated using TreeAnnotator v1.10.4 and viewed on FigTree v1.4.4. Trace files were viewed on Tracer v1.7.1 and all ESS parameters were > 200. The neighbor-joining tree based on SNVs was generated using the R phangorn library ^47^ based on hamming distances calculated using the R 1071 library. All other visualizations and calculations were using R version 4.0.2 (2020-06-22).

### Quantification and Statistical Analysis

Further statistical analysis and visualization were conducted using R v 4.0.0. The code is available on bitbucket.

## Notes

### Competing Interest Statement

The authors have declared no competing interest.

### Clinical Trial

NA

### Author Declarations

Sample use was approved under human subjects approvals # 20-2448 and # 13-2140 by the Institutional Review Board (IRB) at the University of North Carolina; CB 7097, 720 Martin Luther King Jr. Blvd. Bldg # 385, Second Floor, Chapel Hill, NC 27599-7097.

## References

1. Kidd, M., Richter, A., Best, A., Cumley, N., Mirza, J., Percival, B., Mayhew, M., Megram, O., Ashford, F., White, T., et al. (2021). S-variant SARS-CoV-2 lineage B1.1.7 is associated with significantly higher viral loads in samples tested by ThermoFisher TaqPath RT-qPCR. J Infect Dis. 10.1093/infdis/jiab082.

2. Reuken, P.A., Stallmach, A., Pletz, M.W., Brandt, C., Andreas, N., Hahnfeld, S., Loffler, B., Baumgart, S., Kamradt, T., and Bauer, M. (2021). Severe clinical relapse in an immunocompromised host with persistent SARS-CoV-2 infection. Leukemia. 10.1038/s41375-021-01175-8.

3. Choi, B., Choudhary, M.C., Regan, J., Sparks, J.A., Padera, R.F., Qiu, X., Solomon, I.H., Kuo, H.H., Boucau, J., Bowman, K., et al. (2020). Persistence and Evolution of SARS-CoV-2 in an Immunocompromised Host. N Engl J Med 383, 2291–2293. 10.1056/NEJMc2031364.

4. Turner, J.S., Day, A., Alsoussi, W.B., Liu, Z., O’Halloran, J.A., Presti, R.M., Patterson, B.K., Whelan, S.P.J., Ellebedy, A.H., and Mudd, P.A. (2020). SARS-CoV-2 Viral RNA Shedding for More Than 87 Days in an Individual With an Impaired CD8+ T Cell Response. Front Immunol 11, 618402. 10.3389/fimmu.2020.618402.

5. McCarthy, K.R., Rennick, L.J., Nambulli, S., Robinson-McCarthy, L.R., Bain, W.G., Haidar, G., and Duprex, W.P. (2021). Recurrent deletions in the SARS-CoV-2 spike glycoprotein drive antibody escape. Science 371, 1139–1142. 10.1126/science.abf6950.

6. Avanzato, V.A., Matson, M.J., Seifert, S.N., Pryce, R., Williamson, B.N., Anzick, S.L., Barbian, K., Judson, S.D., Fischer, E.R., Martens, C., et al. (2020). Case Study: Prolonged Infectious SARS-CoV-2 Shedding from an Asymptomatic Immunocompromised Individual with Cancer. Cell 183, 1901–1912 e1909. 10.1016/j.cell.2020.10.049.

7. Kemp, S.A., Collier, D.A., Datir, R.P., Ferreira, I., Gayed, S., Jahun, A., Hosmillo, M., Rees-Spear, C., Mlcochova, P., Lumb, I.U., et al. (2021). SARS-CoV-2 evolution during treatment of chronic infection. Nature 592, 277–282. 10.1038/s41586-021-03291-y.

8. McNamara, R.P., Caro-Vegas, C., Landis, J.T., Moorad, R., Pluta, L.J., Eason, A.B., Thompson, C., Bailey, A., Villamor, F.C.S., Lange, P.T., et al. (2020). High-Density Amplicon Sequencing Identifies Community Spread and Ongoing Evolution of SARS-CoV-2 in the Southern United States. Cell Rep 33, 108352. 10.1016/j.celrep.2020.108352.

9. Korber, B., Fischer, W.M., Gnanakaran, S., Yoon, H., Theiler, J., Abfalterer, W., Hengartner, N., Giorgi, E.E., Bhattacharya, T., Foley, B., et al. (2020). Tracking Changes in SARS-CoV-2 Spike: Evidence that D614G Increases Infectivity of the COVID-19 Virus. Cell 182, 812–827 e819. 10.1016/j.cell.2020.06.043.

10. Volz, E., Hill, V., McCrone, J.T., Price, A., Jorgensen, D., O’Toole, A., Southgate, J., Johnson, R., Jackson, B., Nascimento, F.F., et al. (2021). Evaluating the Effects of SARS-CoV-2 Spike Mutation D614G on Transmissibility and Pathogenicity. Cell 184, 64–75 e11. 10.1016/j.cell.2020.11.020.

11. Zhang, W., Davis, B.D., Chen, S.S., Sincuir Martinez, J.M., Plummer, J.T., and Vail, E. (2021). Emergence of a Novel SARS-CoV-2 Variant in Southern California. JAMA. 10.1001/jama.2021.1612.

12. Washington, N.L., Gangavarapu, K., Zeller, M., Bolze, A., Cirulli, E.T., Schiabor Barrett, K.M., Larsen, B.B., Anderson, C., White, S., Cassens, T., et al. (2021). Genomic epidemiology identifies emergence and rapid transmission of SARS-CoV-2 B.1.1.7 in the United States. medRxiv. 10.1101/2021.02.06.21251159.

13. Barzin, A., Schmitz, J.L., Rosin, S., Sirpal, R., Almond, M., Robinette, C., Wells, S., Hudgens, M., Olshan, A., Deen, S., et al. (2020). SARS-CoV-2 Seroprevalence among a Southern U.S. Population Indicates Limited Asymptomatic Spread under Physical Distancing Measures. mBio 11. 10.1128/mBio.02426-20.

14. Danecek, P., Bonfield, J.K., Liddle, J., Marshall, J., Ohan, V., Pollard, M.O., Whitwham, A., Keane, T., McCarthy, S.A., Davies, R.M., and Li, H. (2021). Twelve years of SAMtools and BCFtools. Gigascience 10. 10.1093/gigascience/giab008.

15. Graham, R.L., Becker, M.M., Eckerle, L.D., Bolles, M., Denison, M.R., and Baric, R.S. (2012). A live, impaired-fidelity coronavirus vaccine protects in an aged, immunocompromised mouse model of lethal disease. Nat Med 18, 1820–1826. 10.1038/nm.2972.

16. Young, B.E., Ong, S.W.X., Kalimuddin, S., Low, J.G., Tan, S.Y., Loh, J., Ng, O.T., Marimuthu, K., Ang, L.W., Mak, T.M., et al. (2020). Epidemiologic Features and Clinical Course of Patients Infected With SARS-CoV-2 in Singapore. JAMA 323, 1488–1494. 10.1001/jama.2020.3204.

17. He, X., Lau, E.H.Y., Wu, P., Deng, X., Wang, J., Hao, X., Lau, Y.C., Wong, J.Y., Guan, Y., Tan, X., et al. (2020). Temporal dynamics in viral shedding and transmissibility of COVID-19. Nat Med 26, 672–675. 10.1038/s41591-020-0869-5.

18. Hou, Y.J., Chiba, S., Halfmann, P., Ehre, C., Kuroda, M., Dinnon, K.H., 3rd, Leist, S.R., Schafer, A., Nakajima, N., Takahashi, K., et al. (2020). SARS-CoV-2 D614G variant exhibits efficient replication ex vivo and transmission in vivo. Science 370, 1464–1468. 10.1126/science.abe8499.

19. Plante, J.A., Liu, Y., Liu, J., Xia, H., Johnson, B.A., Lokugamage, K.G., Zhang, X., Muruato, A.E., Zou, J., Fontes-Garfias, C.R., et al. (2021). Spike mutation D614G alters SARS-CoV-2 fitness. Nature 592, 116–121. 10.1038/s41586-020-2895-3.

20. Worobey, M., Pekar, J., Larsen, B.B., Nelson, M.I., Hill, V., Joy, J.B., Rambaut, A., Suchard, M.A., Wertheim, J.O., and Lemey, P. (2020). The emergence of SARS-CoV-2 in Europe and North America. Science 370, 564–570. 10.1126/science.abc8169.

21. Abu-Raddad, L.J., Chemaitelly, H., Malek, J.A., Ahmed, A.A., Mohamoud, Y.A., Younuskunju, S., Al Kanaani, Z., Al Khal, A., Al Kuwari, E., Butt, A.A., et al. (2021). Two prolonged viremic SARS-CoV-2 infections with conserved viral genome for two months. Infect Genet Evol 88, 104684. 10.1016/j.meegid.2020.104684.

22. van Dorp, L., Acman, M., Richard, D., Shaw, L.P., Ford, C.E., Ormond, L., Owen, C.J., Pang, J., Tan, C.C.S., Boshier, F.A.T., et al. (2020). Emergence of genomic diversity and recurrent mutations in SARS-CoV-2. Infect Genet Evol 83, 104351. 10.1016/j.meegid.2020.104351.

23. Zucman, N., Uhel, F., Descamps, D., Roux, D., and Ricard, J.D. (2021). Severe reinfection with South African SARS-CoV-2 variant 501Y.V2: A case report. Clin Infect Dis. 10.1093/cid/ciab129.

24. Vetter, P., Cordey, S., Schibler, M., Vieux, L., Despres, L., Laubscher, F., Andrey, D.O., Martischang, R., Harbarth, S., Cuvelier, C., et al. (2021). Clinical, virological and immunological features of a mild case of SARS-CoV-2 re-infection. Clin Microbiol Infect. 10.1016/j.cmi.2021.02.010.

25. Lee, J.T., Hesse, E.M., Paulin, H.N., Datta, D., Katz, L.S., Talwar, A., Chang, G., Galang, R.R., Harcourt, J.L., Tamin, A., et al. (2021). Clinical and Laboratory Findings in Patients with Potential SARS-CoV-2 Reinfection, May-July 2020. Clin Infect Dis. 10.1093/cid/ciab148.

26. To, K.K., Hung, I.F., Ip, J.D., Chu, A.W., Chan, W.M., Tam, A.R., Fong, C.H., Yuan, S., Tsoi, H.W., Ng, A.C., et al. (2020). COVID-19 re-infection by a phylogenetically distinct SARS-coronavirus-2 strain confirmed by whole genome sequencing. Clin Infect Dis. 10.1093/cid/ciaa1275.

27. Tillett, R.L., Sevinsky, J.R., Hartley, P.D., Kerwin, H., Crawford, N., Gorzalski, A., Laverdure, C., Verma, S.C., Rossetto, C.C., Jackson, D., et al. (2021). Genomic evidence for reinfection with SARS-CoV-2: a case study. Lancet Infect Dis 21, 52–58. 10.1016/S1473-3099(20)30764-7.

28. Van Elslande, J., Vermeersch, P., Vandervoort, K., Wawina-Bokalanga, T., Vanmechelen, B., Wollants, E., Laenen, L., Andre, E., Van Ranst, M., Lagrou, K., and Maes, P. (2020). Symptomatic SARS-CoV-2 reinfection by a phylogenetically distinct strain. Clin Infect Dis. 10.1093/cid/ciaa1330.

29. Lu, S., Xie, X.X., Zhao, L., Wang, B., Zhu, J., Yang, T.R., Yang, G.W., Ji, M., Lv, C.P., Xue, J., et al. (2021). The immunodominant and neutralization linear epitopes for SARS-CoV-2. Cell Rep 34, 108666. 10.1016/j.celrep.2020.108666.

30. Anderson, E.J., Rouphael, N.G., Widge, A.T., Jackson, L.A., Roberts, P.C., Makhene, M., Chappell, J.D., Denison, M.R., Stevens, L.J., Pruijssers, A.J., et al. (2020). Safety and Immunogenicity of SARS-CoV-2 mRNA-1273 Vaccine in Older Adults. N Engl J Med 383, 2427–2438. 10.1056/NEJMoa2028436.

31. Sapoval, N., Mahmoud, M., Jochum, M., Liu, Y., Elworth, R.A.L., Wang, Q., Albin, D., Ogilvie, H., Lee, M.D., Villapol, S., et al. (2021). Hidden genomic diversity of SARS-CoV-2: implications for qRT-PCR diagnostics and transmission. Genome Res. 10.1101/gr.268961.120.

32. Abdul-Jawad, S., Bau, L., Alaguthurai, T., Del Molino Del Barrio, I., Laing, A.G., Hayday, T.S., Monin, L., Munoz-Ruiz, M., McDonald, L., Francos Quijorna, I., et al. (2021). Acute Immune Signatures and Their Legacies in Severe Acute Respiratory Syndrome Coronavirus-2 Infected Cancer Patients. Cancer Cell 39, 257–275 e256. 10.1016/j.ccell.2021.01.001.

33. Zou, L., Ruan, F., Huang, M., Liang, L., Huang, H., Hong, Z., Yu, J., Kang, M., Song, Y., Xia, J., et al. (2020). SARS-CoV-2 Viral Load in Upper Respiratory Specimens of Infected Patients. N Engl J Med 382, 1177–1179. 10.1056/NEJMc2001737.

34. Wajnberg, A., Mansour, M., Leven, E., Bouvier, N.M., Patel, G., Firpo-Betancourt, A., Mendu, R., Jhang, J., Arinsburg, S., Gitman, M., et al. (2020). Humoral response and PCR positivity in patients with COVID-19 in the New York City region, USA: an observational study. Lancet Microbe 1, e283–e289. 10.1016/S2666-5247(20)30120-8.

35. Arons, M.M., Hatfield, K.M., Reddy, S.C., Kimball, A., James, A., Jacobs, J.R., Taylor, J., Spicer, K., Bardossy, A.C., Oakley, L.P., et al. (2020). Presymptomatic SARS-CoV-2 Infections and Transmission in a Skilled Nursing Facility. N Engl J Med 382, 2081–2090. 10.1056/NEJMoa2008457.

36. Wyllie, A.L., Fournier, J., Casanovas-Massana, A., Campbell, M., Tokuyama, M., Vijayakumar, P., Warren, J.L., Geng, B., Muenker, M.C., Moore, A.J., et al. (2020). Saliva or Nasopharyngeal Swab Specimens for Detection of SARS-CoV-2. N Engl J Med 383, 1283–1286. 10.1056/NEJMc2016359.

37. Tomassini, S., Kotecha, D., Bird, P.W., Folwell, A., Biju, S., and Tang, J.W. (2021). Setting the criteria for SARS-CoV-2 reinfection - six possible cases. J Infect 82, 282–327. 10.1016/j.jinf.2020.08.011.

38. Owusu, D., Pomeroy, M.A., Lewis, N.M., Wadhwa, A., Yousaf, A.R., Whitaker, B., Dietrich, E., Hall, A.J., Chu, V., Thornburg, N., et al. (2021). Persistent SARS-CoV-2 RNA Shedding without Evidence of Infectiousness: A Cohort Study of Individuals with COVID-19. J Infect Dis. 10.1093/infdis/jiab107.

39. Lu, J., du Plessis, L., Liu, Z., Hill, V., Kang, M., Lin, H., Sun, J., Francois, S., Kraemer, M.U.G., Faria, N.R., et al. (2020). Genomic Epidemiology of SARS-CoV-2 in Guangdong Province, China. Cell 181, 997–1003 e1009. 10.1016/j.cell.2020.04.023.

40. Ramirez, F., Ryan, D.P., Gruning, B., Bhardwaj, V., Kilpert, F., Richter, A.S., Heyne, S., Dundar, F., and Manke, T. (2016). deepTools2: a next generation web server for deep-sequencing data analysis. Nucleic Acids Res 44, W160–165. 10.1093/nar/gkw257.

41. Rambaut, A., Holmes, E.C., O’Toole, A., Hill, V., McCrone, J.T., Ruis, C., du Plessis, L., and Pybus, O.G. (2020). A dynamic nomenclature proposal for SARS-CoV-2 lineages to assist genomic epidemiology. Nat Microbiol 5, 1403–1407. 10.1038/s41564-020-0770-5.

42. Coronaviridae Study Group of the International Committee on Taxonomy of, V. (2020). The species Severe acute respiratory syndrome-related coronavirus: classifying 2019-nCoV and naming it SARS-CoV-2. Nat Microbiol 5, 536–544. 10.1038/s41564-020-0695-z.

43. Katoh, K., and Standley, D.M. (2013). MAFFT multiple sequence alignment software version 7: improvements in performance and usability. Mol Biol Evol 30, 772–780. 10.1093/molbev/mst010.

44. Ronquist, F., Teslenko, M., van der Mark, P., Ayres, D.L., Darling, A., Hohna, S., Larget, B., Liu, L., Suchard, M.A., and Huelsenbeck, J.P. (2012). MrBayes 3.2: efficient Bayesian phylogenetic inference and model choice across a large model space. Syst Biol 61, 539–542. 10.1093/sysbio/sys029.

45. Stamatakis, A. (2014). RAxML version 8: a tool for phylogenetic analysis and post-analysis of large phylogenies. Bioinformatics 30, 1312–1313. 10.1093/bioinformatics/btu033.

46. Bouckaert, R., Vaughan, T.G., Barido-Sottani, J., Duchene, S., Fourment, M., Gavryushkina, A., Heled, J., Jones, G., Kuhnert, D., De Maio, N., et al. (2019). BEAST 2.5: An advanced software platform for Bayesian evolutionary analysis. PLoS Comput Biol 15, e1006650. 10.1371/journal.pcbi.1006650.

47. Schliep, K.P. (2011). phangorn: phylogenetic analysis in R. Bioinformatics 27, 592–593. 10.1093/bioinformatics/btq706.

